# Acutelines: a Dutch emergency department-based data-biobank for multimodal, longitudinal acute care research

**DOI:** 10.64898/2026.07.30.26359325

**Authors:** Raymond J. van Wijk, Juan Miguel Lopez Alcaraz, Anna D. Schoonhoven, Jie Li, Sanne Ter Horst, Michèle A. ter Voert, Annet M. Eerens, Anna J. la Bastide, Jochem Postema, Jan C. ter Maaten, Ewoud ter Avest, Nils Strodthoff, Hjalmar R. Bouma

## Abstract

**Background:** High-quality, longitudinal data from the Emergency Department (ED) are essential for understanding acute illness trajectories and for developing clinically deployable prediction models. However, most existing ED datasets are static, limited in modality or disconnected from long-term outcomes or biomaterials.

**Methods:** We describe a two-year sample of Acutelines, a continuously operating ED-based data-biobank infrastructure at the University Medical Center Groningen. Acutelines is embedded in routine care and captures the full acute care trajectory and uses a stepped consent procedure. It integrates demographics, vital signs, laboratory results, diagnoses, treatments, waveforms, patient-reported outcomes, and prospectively collected biomaterials. Data are linked to in-hospital outcomes and post-discharge mortality.

**Results:** The presented dataset comprises 29,314 ED visits from 18,850 adult patients. Clinical variables are available for nearly all visits, with additional data and questionnaires (n=4,963) and biomaterials (n=2,723) collected in predefined subgroups. Associations between early triage features (National Early Warning Score, vital signs, diagnostic categories) and outcomes, illustrate the clinical depth and longitudinal value of the infrastructure.

**Conclusion:** Acutelines is a living, continuously expanding research infrastructure. Its combination of ED-first inclusion, multimodal data, linkage to long-term outcomes, and availability of biomaterials distinguishes it from existing datasets. Acutelines provides a robust foundation for hypothesis generation, translational research, and the development, validation, and benchmarking of data-driven clinical decision support tools and biomarker tests in acute medicine, while its ongoing expansion enables prospective studies and interventional research.

**Trial registration:** Acutelines is registered at ClinicalTrials.gov under trial registration number NCT04615065.

## Background

Clinical decision making in the emergency department (ED) is central to acute care, determining which patients require immediate care, how clinical resources are allocated and determining the level of care needed after the stay at the ED under conditions of uncertainty and time pressure [1]. Decisions made at ED presentation influence not only immediate treatment but also later clinical courses, including hospital admission, escalation of care, and long-term outcomes, as well as the burden on the healthcare system [2,3]. Acute care demands continue to rise globally due to aging populations, multimorbidity, leading to increased strain on acute care. Therefore, efforts to support efficient, accurate clinical decision making in acute care have become a major public health priority [4]. However, high-quality datasets that capture real-world ED practice with sufficient breadth, temporal resolution, and follow-up remain scarce. Many existing acute care datasets are disease-specific, retrospectively assembled, or based on time-limited inclusion periods, often selecting patients based on final diagnoses or outcomes rather than initial ED presentation [5]. Such approaches risk selection bias and fail to reflect the heterogeneity and uncertainty inherent to early acute care decision making. Although complementary datasets containing structured clinical data, ED encounters, and high-resolution physiological signals have been introduced in recent years, they remain few in number and provide only partial coverage across clinical modalities, additional biomaterial collection, and longitudinal follow-up [6,7].

To address these limitations, the University Medical Center Groningen (UMCG) founded Acutelines as an integrated research infrastructure, comprising dedicated research staff embedded in the ED and a linked data-biobank [8]. The Netherlands, with its structured and accessible healthcare system, provides a unique opportunity to examine triage practices in a high-resource setting [9]. The UMCG serves as one of the largest tertiary and transplant referral centers in the Netherlands, providing academic acute care for a large and diverse geographical catchment area that includes both urban and rural populations and functions in close interaction with regional general hospitals. As a result, the UMCG ED receives a broad and heterogeneous spectrum of patients with acute diseases, supporting its relevance as a representative setting for emergency care research in a high-income country. Acutelines captures the complete acute care trajectory, from prehospital information and emergency department triage through in-hospital treatment and longitudinal follow-up, including ward and ICU admissions and all-cause mortality. The cohort integrates routinely collected clinical data with extended clinical scores, adjudicated diagnostic information, continuously acquired bedside monitoring and high-resolution physiological waveforms, patient-reported outcome measures, and prospectively collected biomaterials. Data are obtained through direct linkage with the electronic health record and external registries, complemented by manual curation and a dedicated biobank. Custom-built software supports real-time patient screening and efficient recruitment within routine emergency department workflows. Using design principles from clinical informatics and biomedical engineering, Acutelines enables continuous data capture with minimal disruption to clinical practice, supporting both prospective and retrospective clinical research. A stepped consent procedure enables inclusion of acutely ill patients while safeguarding autonomy and minimizing consent-related selection bias [8,10].

This cohort provides a robust foundation for data-driven research aimed at improving acute care outcomes. Its relevance spans multiple research domains, including early identification of patients at risk of clinical deterioration using high-resolution electrophysiological waveforms and clinical data [11–13], development of machine learning (ML) models at ED triage across diverse diagnostic and deterioration scenarios [14], and predictive enrichment strategies to identify patients most likely to benefit from specific therapies [15]. In addition, Acutelines provides a flexible platform for embedding dedicated prospective studies and interventional trials within routine care. By detailing the construction, scope, and governance of Acutelines, and by presenting a detailed two-year sample of the cohort (n = 29,314 ED visits), we aim to accelerate hypothesis generation, model development, and translational research in emergency medicine across Europe, and to stimulate collaborative use of this infrastructure. Although data and biomaterials are not publicly accessible due to privacy and ethical constraints, Acutelines actively supports collaborative research through regulated access procedures. By fostering cross-institutional collaboration, this infrastructure enables reproducible research, benchmarking and external validation of clinical artificial intelligence (AI) tools, and healthcare innovation aligned with the principles of learning health systems.

## Methods

### Study design

Acutelines is a multidisciplinary, prospective, continuously operating data-biobank infrastructure embedded in routine ED care at the UMCG [8]. It was designed to enable systematic reuse of routine clinical data and targeted collection of extended data and biomaterials in acutely ill patients presenting at the ED.

### Population and inclusion

Adult patients presenting to the ED are screened for inclusion based on predefined criteria reflecting acuity and target diagnoses. Routine clinical data are reused by opt-out, while extended de novo data and biomaterials are collected in predefined high-acuity and diagnosis-specific subgroups, as detailed below. Adult patients triaged with an Emergency Severity Index (ESI) level 1 to 3 (red, orange, or yellow) are automatically screened in the electronic health record for a registered objection to research and, if eligible, are automatically associated with Acutelines. This enables systematic capture of routine clinical data from the electronic health record and bedside monitoring systems, including all vital parameters and high-resolution electrophysiological waveforms. A graphical overview of the included data is shown in Figure 1.

**Figure 1:**
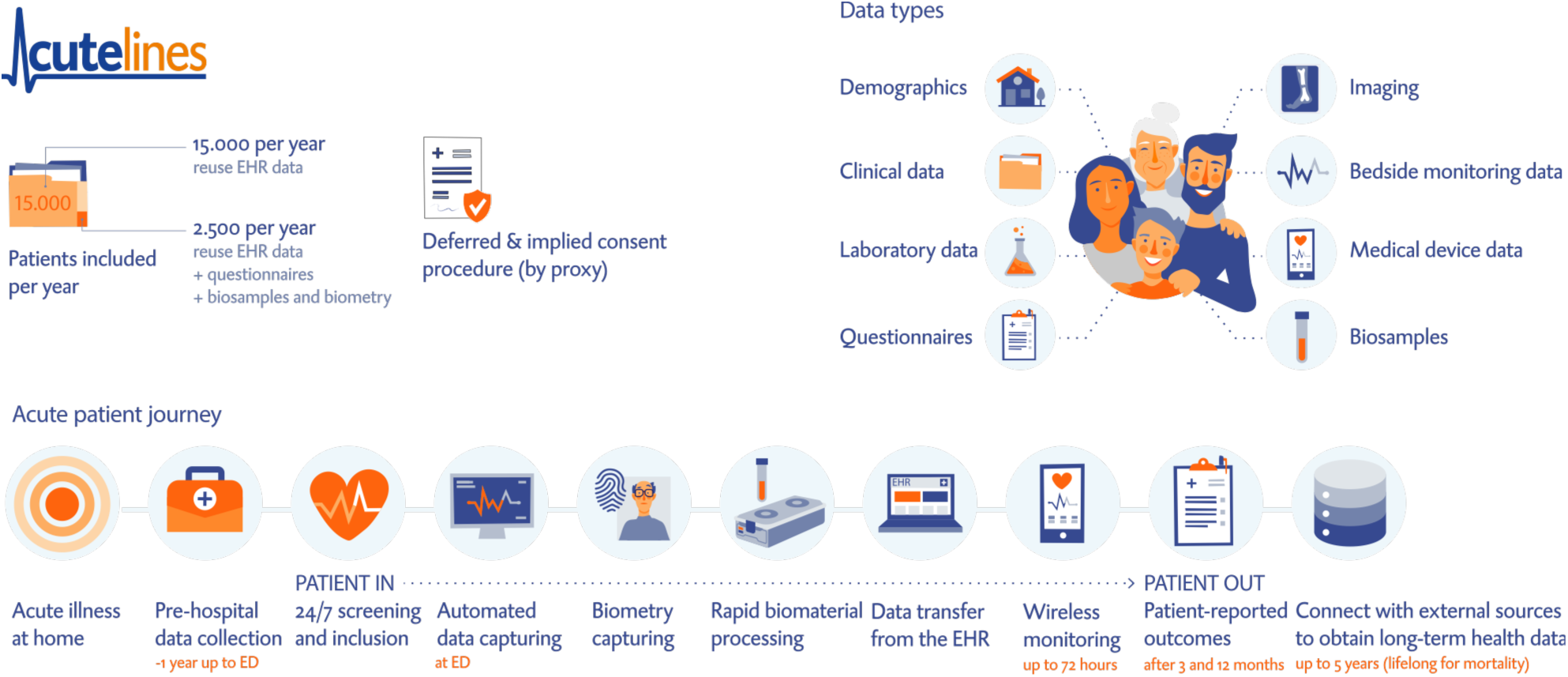
Schematic overview of data collection within the Acutelines infrastructure. Approximately 15,000 patient visits are included per year. The acute patient journey is represented from the start of acute illness onset at home through pre-hospital data collection (up to one year before ED admission, including ambulance data), emergency department admission, and automated capture of clinical data. This includes biometry acquisition, rapid processing of biomaterials, transfer of data from the electronic health record, bedside monitors and, when eligible, wireless monitoring of vital signs during hospitalization. Following discharge, short– and long-term outcomes are linked through hospital records and external registries, enabling follow-up for up to 5 years or until death. Collected data modalities comprise demographic information, clinical and laboratory data, questionnaires, imaging, bedside monitoring and medical device data, and biospecimens.

In addition, patients presenting to the ED for internal medicine (and subspecialities), gastro-enterology, pulmonology, rheumatology, or emergency medicine with high triage acuity (ESI 1–2), medium triage acuity (ESI 3) combined with ambulance or helicopter transport or presenting with predefined target diagnoses (a.o., infection/sepsis, shock, syncope, anaphylaxis, intoxications, asthma/COPD exacerbations, venous thromboembolism, and metabolic emergencies) are eligible for collection of extended de novo data and biomaterials. These data and samples are obtained between 05.00 and 23.00 by the ED nurse supported by trained research assistants. Outside this timeslot, routine clinical data are continuously collected through opt-out.

### Consent and ethical framework

A stepped, adapted consent procedure is implemented to balance respect for patient autonomy with feasibility in acute care and minimization of consent-related selection bias [10]. Routine clinical data are included unless patients have registered an objection (opt-out). For de novo data and biomaterial collection written informed consent is obtained whenever possible, deferred consent (by proxy), or in specific circumstances (e.g., death before consent can be obtained or failure to obtain consent within 30 days) by opt-out, is sought when patients are temporarily incapacitated. Participants can withdraw their participation (either consent or automatic inclusion) in the cohort at any time without providing a reason.

All procedures are approved by the institutional review board (#201900635) and comply with national and European regulations on medical research and data protection. Data and biomaterials from Acutelines are available to researchers in collaborative research projects and after ethical approval of the study protocol by the UMCG institutional review board. The current Acutelines’ resource description was approved by the ethics committee of the UMCG (#23514). The Strengthening the Reporting of Observational Studies in Epidemiology (STROBE) guidelines were followed.

### Registration and transparency

Acutelines is registered at ClinicalTrials.gov under trial registration number NCT04615065. Information for participants is publicly available at acutelines.umcg.nl, while detailed information for researchers, including governance procedures and data access, is provided at www.acutelines.nl. Acutelines is also listed in the UMCG Research Data Catalogue at umcgresearchdatacatalogue.nl, where the scope, structure, and reuse conditions of the infrastructure are described to support transparency and discoverability. Detailed information on the Acutelines cohort and participant selection has been published previously [8]. The infrastructure is continuously reviewed and refined to align with evolving research questions and methodological advances; the present manuscript describes the initial design and its current implementation.

## Data sources and collection

### Re-use of clinical data

Acutelines systematically captures routine clinical data through direct linkage with the electronic health record (EPIC), including demographics, triage characteristics, vital signs, laboratory results, imaging, treatments, diagnoses, and outcomes. Continuous bedside monitoring data are acquired directly from ED monitoring systems and include both vital parameters (1 Hz; heart rate, oxygen saturation, respiratory rate) and high-resolution physiological waveforms, measured at 500 Hz for electrocardiography and 125 Hz for photoplethysmography and thoracic impedance. Data are captured from Philips IntelliVue monitors via Capsule Neuron 3, temporarily buffered in a proprietary waveform environment, and transferred to permanent storage on a daily basis with indexed metadata to enable efficient retrieval. All data streams are integrated within the institutional data warehouse and complemented by linkage to external registries for follow-up and outcome ascertainment, after which they are made available for extraction for patients included in Acutelines.

### De novo data & biomaterials

Additional de novo data are collected by trained research staff and comprise structured clinical assessments, adjudicated diagnostic information, and validated patient-reported outcome measures (PROMs). Data elements include extended demographic characteristics (a.o., marital status, educational level), lifestyle factors (a.o., smoking, alcohol and drug use, activity, nutrition), functional status, cognitive screening instruments (a.o., DOSS, 4-AT), wearable data, face photographs, and structured free-text fields from nursing and physician assessments. De novo data are collected using REDCap (Research Electronic Data Capture), a secure, web-based platform designed to support validated research data capture, including audit trails, automated export procedures, and interoperability with external sources [16,17]. Facial photographs are stored separately in secure institutional file storage.

#### Biomaterials

In parallel, Acutelines maintains a biobank with prospectively collected biological samples obtained at ED presentation with follow-up sampling during early hospitalization according to standardized protocols. Biomaterials are collected in predefined subgroups eligible for de novo data collection, including patients with high triage acuity (ESI 1-2) and those presenting with a suspected infection or sepsis, shock, or related acute conditions. When eligible, samples are obtained directly in the emergency department, with additional follow-up sampling during hospitalization. All samples are processed within 4 hours and stored at –80 °C. Sample handling and metadata, including time to centrifugation, time to freezing, freeze–thaw cycles, and processing events, are systematically recorded using OpenSpecimen (Krishagni Solutions, India).

#### Follow-up questionnaires

Patients included for de novo data collection who were subsequently hospitalized are invited to complete follow-up questionnaires at three months and one year after admission. These questionnaires capture PROMs, functional status, physical activity, and changes in demographic characteristics or lifestyle factors, enabling longitudinal assessment of health status beyond the acute episode.

#### Outcome and follow-up

Captured outcomes for all patients include ED disposition, hospital and ICU admission, length of stay, readmission, and short– and long-term mortality. For patients included with de novo data collection, these outcomes are complemented by patient-reported data from follow-up questionnaires and can be linked to external registries, including general practice records, national population and mortality registries, and health care insurance databases.

### Data curation and quality control

Automated data extraction is complemented by manual curation by trained research staff. Clinical scores extending beyond routine care are derived, and diagnostic information is adjudicated using standardized definitions and recorded in REDCap. Diagnoses and comorbidities are coded both as free text and using International Classification of Diseases, 10th Revision (ICD-10) and SNOMED-CT in routine care by clinical staff within EPIC, and are supplemented by additional adjudication in REDCap by research staff. Data processing and quality control are supported by dedicated in-house software, including the Acutelines research assistant helper application (Raha) for screening and recruitment, and custom-built dashboards using Grafana (Grafana Labs, New York, NY, USA) to monitor inclusion rates and data completeness. Data collected via REDCap are integrated with EPIC electronic health record data within the UMCG data warehouse, resulting in a uniform, linked dataset. This integrated approach enables consistent, high-quality longitudinal and cross-sectional analyses across multiple data sources, supporting both clinical and data-driven research in acute care. To facilitate preprocessing and cleaning of exported datasets, we developed the Acutelines datatoolbox [18], an open-source R package that supports standardized handling of Acutelines data extracts and enables common post hoc calculations while preserving transparency of clinical assumptions and methodological choices.

### Data access

Data and biomaterials are available for collaborative research through regulated access procedures following protocol approval, ensuring appropriate governance and data protection.

### Data analysis

For the current data resource paper, we selected a two-year sample of all patients included in Acutelines. After data extraction, preprocessing, and cleaning using the Acutelines datatoolbox (version 1.1.0) [18], laboratory parameters were harmonized and post hoc clinical scores were derived, including Sequential Organ Failure Assessment (SOFA) and National Early Warning Score (NEWS) [19–21]. ICD-10 codes were aggregated at the ICD-10 block level to improve interpretability and retain clinically relevant diagnostic clusters [22]. We report baseline demographic, triage, laboratory, and outcome characteristics of emergency department visits in this two-year sample (N=29,314). In addition, we describe the associations between the 20 most frequent diagnostic categories, initial severity at triage (i.e., NEWS and vital parameters) and critical outcomes including hospital admission, ICU admission (including in-hospital deterioration), readmission, and mortality. Supplementary analyses provide detailed distributions of demographics, logistics, mortality, vital signs, laboratory parameters, SOFA scores during the first 72 hours, common diagnoses and comorbidities, as well as the availability of de novo data and biomaterials.

## Results

### Descriptive statistics

Between November 1, 2023 and November 1, 2025, Acutelines included 18,850 unique adult patients, accounting for 29,314 emergency department visits. Based on eligibility, absence of objection by the participant, and presentation during capture hours (05.00– 23.00, seven days per week), 4,963 visits (17%) were enriched with extended de novo data, including structured assessments and follow-up questionnaires. Biomaterials were collected during 2,723 patient visits (Table 1, Supplementary Table A.1). High-resolution time-series and electrophysiological waveform data were collected for a substantial subset of visits, with detailed availability summarized in Supplementary Table A.2.

**Table 1:** Baseline demographic, triage, laboratory, and outcome characteristics of emergency department visits in the Acutelines cohort (N = 2S,314). Values are reported as counts with percentages or medians with interquartile ranges (Q1–Q3), based on data available at emergency department presentation or during subsequent hospitalisation. Sample sizes for each variable are indicated due to variable data availability.

| Characteristic | Data available (N) | Values<sup>†</sup> |
| --- | --- | --- |
| **Baseline & demographics** |  |  |
| Sex | 29,314 |  |
| - Female | 13,058 | (45%) |
| - Male | 16,256 | (55%) |
| Age (years) | 29,314 | 62 (45, 73) |
| Number of Comorbidities | 29,314 | 5 (1, 12) |
| **Triage measurements** |  |  |
| Triage ESI | 28,785 |  |
| - 1 (Red) |  | 929 (3.2%) |
| - 2 (Orange) |  | 6,378 (22%) |
| - 3 (Yellow) |  | 20,420 (71%) |
| - 4 (Green) |  | 960 (3.3%) |
| - 5 (Blue) |  | 98 (0.3%) |
| Heart Rate (/min) | 26,849 | 85 (72, 99) |
| Temperature (°C) | 23,616 | 36.70 (36.10, 36.70) |
| Respiratory Rate (/min) | 21,195 | 16 (14, 20) |
| Oxygen Supplementation at arrival | 29,314 | 1,934 (6.6%) |
| Oxygen saturation (%) | 26,892 | 97 (95, 99) |
| Systolic blood pressure (mmHg) | 26,580 | 136 (121, 153) |
| Diastolic blood pressure (mmHg) | 26,580 | 82 (72, 92) |
| Mean Arterial Pressure (mmHg) | 26,580 | 100 (90, 112) |
| GCS | 6,238 | 15 (15, 15) |
| AVPU | 2,322 |  |
| - Alert |  | 2,257 (97%) |
| - Other (Verbal, Pain, Unresponsive) |  | 65 (3%) |
| NEWS | 27,455 | 1 (0, 3) |
| **Laboratory measurements** |  |  |
| Hemoglobin (mmol/L) | 25,506 | 8.10 (7.10, 8.90) |
| Platelets (<sup>9</sup>/L) | 25,449 | 242 (189, 306) |
| WBC (<sup>9</sup>/L) | 25,471 | 9.1 (6.9, 12.2) |
| Erythrocytes (<sup>12</sup>/L) | 14,884 | 4.31 (3.76, 4.76) |
| INR | 6,875 | 1.10 (1.00, 1.40) |
| Glucose (mmol/L) | 22,315 | 6.60 (5.70, 8.20) |
| Sodium (mmol/L) | 24,520 | 138 (135, 140) |
| Potassium (mmol/L) | 24,567 | 4.10 (3.80, 4.50) |
| Creatinine (mmol/L) | 24,800 | 79 (64, 103) |
| Calcium (mmol/L) | 17,952 | 2.32 (2.23, 2.39) |
| Blood Urea Nitrogen (mmol/L) | 24,578 | 6.2 (4.6, 8.5) |
| Total Bilirubin (umol/L) | 19,365 | 8 (5, 12) |
| Direct Bilirubin (umol/L) | 2,676 | 13 (10, 22) |
| Albumin (g/L) | 16,366 | 40 (36, 43) |
| ASAT (U/L) | 22,906 | 27 (21, 38) |
| ALAT (U/L) | 22,948 | 20 (13, 32) |
| CRP (mg/L) | 24,359 | 8 (2, 45) |
| Lactate (mmol/L) | 17,982 | 1.40 (1.00, 2.00) |
| eGFR (mL/min/1.73m<sup>2</sup>) | 24,623 | 82 (58, 98) |
| Arterial pO<sub>2</sub> (kPa) | 4,676 | 10 (8, 14) |
| Arterial pCO<sub>2</sub> (kPa) | 4,678 | 5.20 (4.50, 6.10) |
| **Logistics & outcome** |  |  |
| De Novo data collected | 4,963 | (17%) |
| ED Length of Stay (hours) | 29,314 | 3.6 (2.4, 5.7) |
| Time till ICU Admission (hours) | 1,849 | 1 (0, 15) |
| ICU Length of Stay (hours) | 1,848 | 41 (21, 92) |
| Ward Length of Stay (hours) | 14,498 | 96 (44, 200) |
| Time till Readmission (days) | 10,597 | 45 (12, 139) |
| Time till Death (days) | 4,435 | 67 (16, 203) |
| Deceased in Hospital | 29,314 | 794 (2.7%) |

<sup>†</sup>n (%); Median (Q1, Q3) Abbreviations: ESI: Emergency Severity Index, GCS: Glasgow Coma Scale, AVPU: Alert, Verbal, Pain, Unresponsive, NEWS: National Early Warning Score, WBC: White Blood Cell count, INR: International Normalized Ratio, ASAT: Aspartate transaminase, ALAT: Alanine transaminase, eGFR: Estimated Glomerular Filtration Rate, pO<sub>2</sub>: Partial oxygen pressure, pCO<sub>2</sub>: Partial carbon dioxide pressure, ED: Emergency Department, ICU: Intensive Care Unit.

### Baseline characteristics

The median age at presentation was 62 years (IQR 45–73), with a slight predominance of male patients (55%) (Table 1; Supplementary Figure A.3). The cohort had a substantial comorbidity burden, with a median of five comorbid conditions per visit (Table 1; Supplementary Figure A.4). The most prevalent ICD-10 comorbidity chapters, as coded by the clinical staff, were diseases of the circulatory system (10%), symptoms not otherwise specified (9%), factors influencing health status (9%), musculoskeletal disorders (8%), neoplasms (8%), and diseases of the digestive system (7%) (Supplementary Figure A.4). The most common emergency department diagnoses, based on ICD-10 labels registered by clinical staff, were injury and poisoning (20%), symptoms not otherwise specified (16%), circulatory diseases (16%), respiratory diseases (7%), digestive diseases (6%), and factors influencing health status (5%) (Supplementary Figure A.5).

At presentation, for most patients vital signs were within normal ranges, with a median NEWS of 1 (IQR 0–3), although marked interindividual variability was observed (Table 1; Supplementary Figures A.6 and A.7). Most visits were triaged as ESI level 3 (yellow, approximately 70%), followed by level 2 (orange, approximately 20%); fewer than 5% were classified as level 1 (red), with similarly small proportions classified as levels 4 or 5 (green and blue) (Supplementary Figure A.8). Laboratory data encompassed a broad range of routine hematological, biochemical, renal, hepatic, and inflammatory parameters, with availability reflecting clinical indication (Table 1). Detailed distributions of laboratory values are shown in Supplementary Figures A.9–A.12.

Patients attended the ED predominantly during weekdays, with peaks during daytime and early evening hours (Supplementary Figure A.8). Most patients presented from home (approximately 90%) and were also discharged home after hospitalization (Supplementary Figure A.8). Median emergency department length of stay was 3.6 hours (IQR 2.4–5.7) (Table 1). Among patients admitted to the intensive care unit, time from emergency department presentation to ICU admission was 1 hour (IQR 0–15), with a median ICU length of stay of 41 hours (IQR 21–92) (Table 1; Supplementary Table A.8). For nursingward admissions, median length of stay was 96 hours (IQR 44–200) (Table 1; Supplementary Figure A.8). Among patients with readmission, median time to readmission was 45 days (IQR 12–139) (Table 1; Supplementary Figure A.13).

Overall, in-hospital mortality was 2.7%, and linkage to longitudinal data enabled assessment of long-term mortality after ED presentation. Among deceased patients, median time to death was 67 days (IQR 16–203) (Table 1; Supplementary Figures A.13 and A.14).

### Outcome stratified by diagnosis– or presenting complaint

The 20 most frequent ICD-10 diagnosis based clusters (ICD-10 blocks [22]) together accounted for approximately two-thirds of all emergency department visits in the two-year sample (Figure 2, Supplementary Figures B.1 and A.5), underscoring that a limited number of diagnostic categories capture the majority of acute presentations. Injury-related diagnosis based clusters were most prevalent, with head injury and injuries of the trunk or extremities together representing roughly 25% of all visits. These categories were associated with high hospital admission rates, commonly exceeding 50%, but relatively low short-term and one-year mortality, generally below 5%, reflecting high acute care utilization with favorable long-term outcomes.

**Figure 2:**
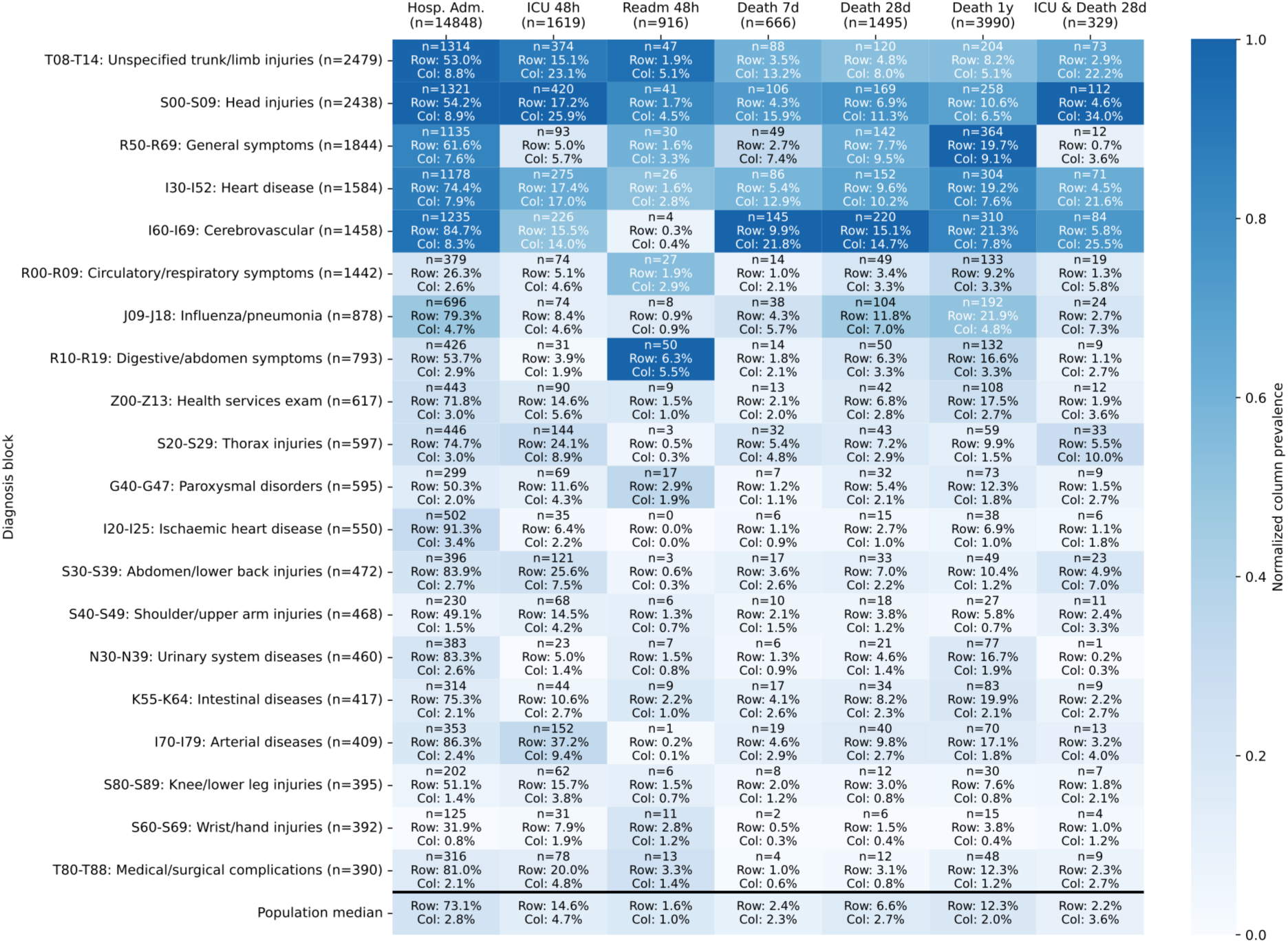
Top 20 ICD-10 clusters stratified by clinical outcomes relative to emergency department presentation. Outcomes include hospital admission, ICU admission within 48 hours, readmission within 48 hours, and mortality at 7 days, 28 days, 1 year, and ICU or death within 28 days. Each cell reports absolute counts together with row– and column-level percentages. Color intensity reflects column-wise normalized prevalence, with darker shades indicating a higher prevalence of a given diagnosis cluster relative to other diagnoses within the same outcome. Hosp. Adm.: Hospital Admission; ICU: Intensive Care Unit; Readm: Readmission.

In contrast, diagnostic clusters related to diseases of the circulatory system, cerebrovascular disease, and non-specific symptoms and abnormal clinical and laboratory findings comprised a smaller fraction of visits, each accounting for less than 10% individually, yet contributed disproportionately to adverse outcomes. For cerebrovascular disease, hospital admission rates exceeded 80%, ICU admission was frequent, and one-year mortality was above 20%, marking this category as one of the highest-risk groups despite its lower prevalence.

Complaint-based clusters, including non-specific complaints and abnormal clinical findings, displayed marked outcome heterogeneity. While a substantial proportion of these visits resulted in rapid discharge, a non-negligible subset was associated with high rates of hospital admission and one-year mortality exceeding 15%, illustrating the diagnostic uncertainty and risk concentration inherent to these early ED presentations. Together, these findings demonstrate that high-level diagnostic grouping at ED presentation already stratifies patients along clinically meaningful gradients of resource use and short– and long-term risk.

### Outcome stratified by vital signs

Abnormal vital parameters at triage were relatively uncommon in absolute terms but showed strong associations with adverse outcomes (Figure 3, Supplementary Figures B.2 and A.6). Hypotension, tachypnea, hypoxia, and altered mental status each occurred in fewer than 10% of visits. Vital parameters at presentation demonstrated clear and clinically interpretable gradients in downstream risk.

**Figure 3:**
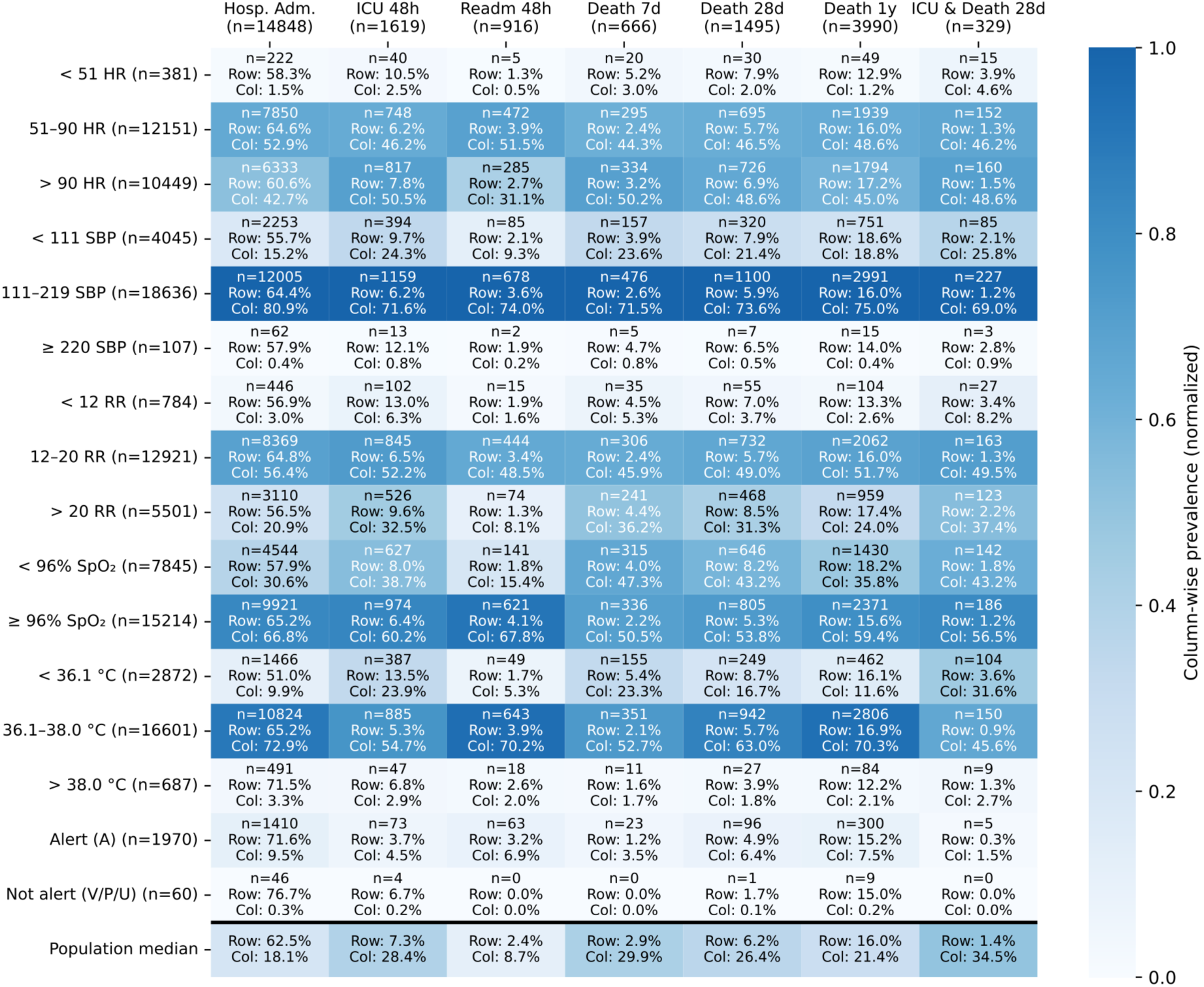
Vital parameters at triage stratified by clinical outcomes relative to emergency department presentation. Outcomes include hospital admission, ICU admission within 48 hours, readmission within 48 hours, and mortality at 7 days, 28 days, 1 year, and ICU or death within 28 days. Each cell displays both row– and column-level prevalence. Color intensity reflects column-wise normalized prevalence, with darker shades indicating a higher prevalence of a given vital parameter category relative to other categories within the same outcome. Hosp. Adm.: Hospital Admission; ICU: Intensive Care Unit; Readm: Readmission; HR: Heart rate; RR: respiratory rate; SpO_2_: peripheral oxygen saturation; SBP: Systolic Blood Pressure; °C: degree Celsius; A: Alert; V: Verbal; P: Pain; U: Unresponsive.

Patients with deviating respiratory vital parameters (SpO_2_<96% or respiratory rate >20/min) had higher ICU admission rates and higher one-year mortality compared to patients with normal reporatoiry vital signs (SpO_2_≥96% or respiratory rate ≥20/min).

Systolic blood pressure (SBP) and heart rate (HR) further stratified outcomes. A higher HR (>90/min) or lower SBP (<111 mmHg) showed a higher incidence of ICU admission and 1-year mortality compared to normal HR and SBP. While a lower HR (<51/min) or higher SBP (≥220 mmHg) was associated with fewer hospital admissions and 1-year mortality, but more ICU admissions, when compared to normal.

Temperature categories demonstrated complementary risk profiles. Patients with normal temperatures had hospital admission rates of 65.2%, ICU admission of 5.3%, and one-year mortality of 16.9%. Fever (≥38.0 °C) was associated with higher hospital admission (71.5%) and ICU admission (6.8%) but lower one-year mortality (12.2%), whereas hypothermia (<36.1 °C) showed lower hospital admission (51.0%) but higher ICU admission (13.5%) and 16.1% one-year mortality.

Together, these findings show that deviations from normal vital parameters are strongly associated with adverse outcomes, while values within normal ranges do not exclude severe trajectories, underscoring the added value of granular vital sign data and novel tools beyond composite scores and classic vital signs.

### Outcome stratified by triage severity

The NEWS score ranges from 0 to 20, with commonly used cut-offs for low (0–4), medium (4–7), and high (≥7) risk of clinical deterioration. NEWS at triage was low for most visits, with approximately half of visits presenting with NEWS 0–1 and fewer than 10% with NEWS ≥ 5 (Figure 4, Supplementary Figures B.3 and A.7). Despite their lower prevalence, higher NEWS categories accounted for a substantial share of critical outcomes. Hospital admission and one-year mortality increased progressively with higher NEWS: admission rates rose from 42.2% for NEWS 0 to 75.5% for NEWS 6, while one-year mortality increased from 9.1% to 26.2% over the same range. Although patients with NEWS ≥5 respresented a minority of ED visits, they accounted for a disproportionate share of ICU admissions and deaths.

**Figure 4:**
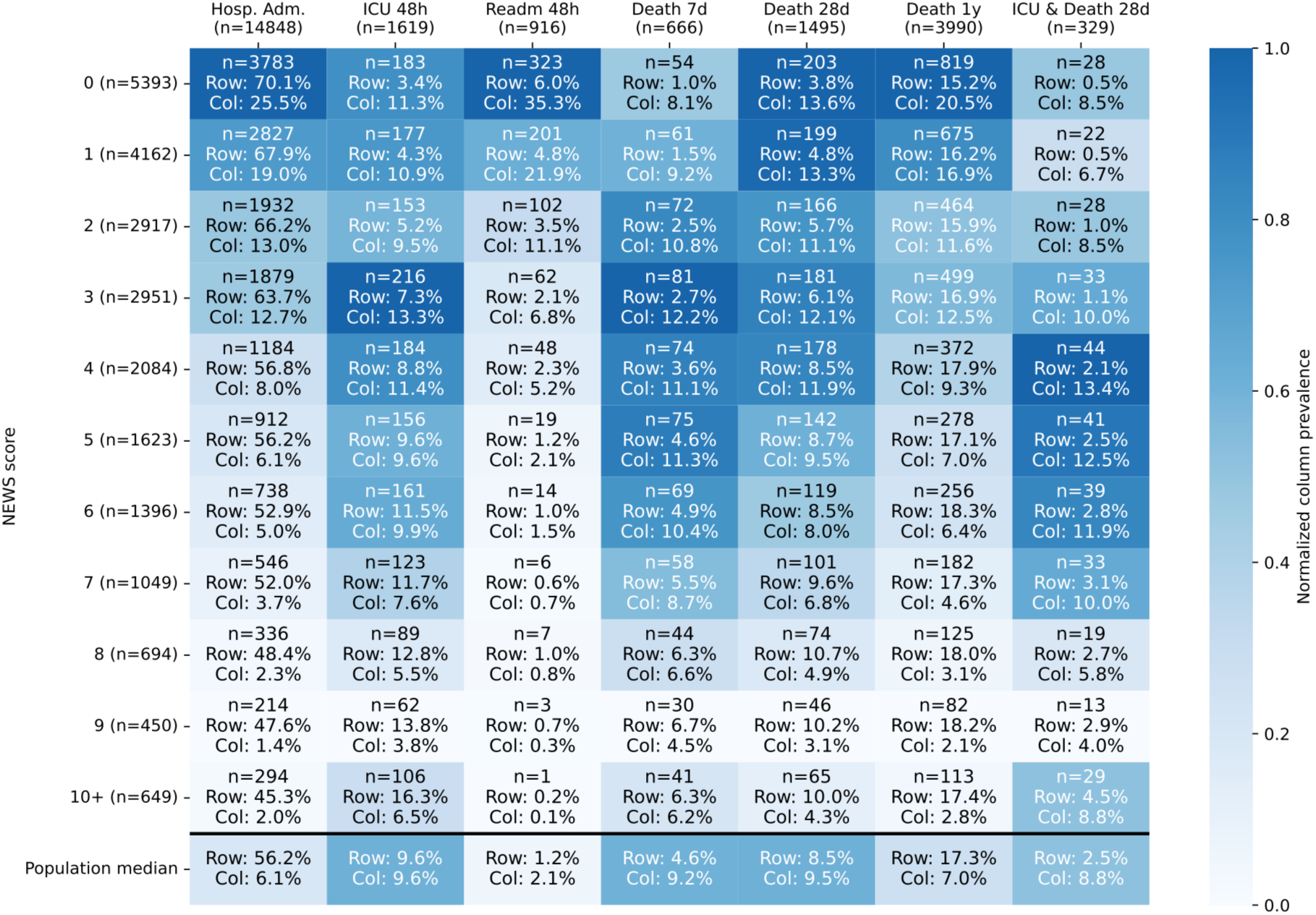
National Early Warning Score (NEWS) at triage stratified by clinical outcomes relative to emergency department presentation. Outcomes include hospital admission, ICU admission within 48 hours, readmission within 48 hours, and mortality at 7 days, 28 days, 1 year, and ICU or death within 28 days. Each cell displays both row– and column-level prevalence. Color intensity reflects column-wise normalized prevalence, with darker shades indicating a higher prevalence of a given NEWS category relative to other categories within the same outcome. Hosp. Adm.: Hospital Admission; ICU: Intensive Care Unit; Readm: Readmission; NEWS: National Early Warning Score.

## Discussion

This data resource paper describes Acutelines as a continuously operating ED based data-biobank that combines routinely collected clinical data with extended *de novo assessments*, electrophysiological waveforms, prospectively collected biomaterials and follow-up questionnaires. The two-year sample demonstrates substantial scale (with 18,850 unique patients and 29,314 emergency department visits), and significant heterogeneity with a broad range in disease severities (ESI), complaints– and diagnosis being included. In addition, it shows clinically meaningful gradients in outcomes across diagnostic and physiological strata. These characteristics position Acutelines as a pragmatic infrastructure for epidemiological studies and for the development, validation, and benchmarking of prediction models and biomarker-based diagnostics. Furthermore, it serves as a robust platform for prospective studies by leveraging on-premise research assistants and an integrated consent framework.

### The challenge of risk discrimination in low– and intermediate-acuity ED patients

A central observation is that information available at ED presentation already captures strong and clinically meaningful risk gradients. Importantly, adverse outcomes were not confined to a small, easily identifiable high-risk subgroup. Trauma-related diagnoses were predominantly associated with hospital admission and short-term ICU care, whereas presentations for non-trauma medical reasons were more frequently linked to longer-term mortality, a pattern consistent with other Dutch emergency department populations [23]. Although low National Early Warning Score values (0–3) were common, a large absolute number of admissions and deaths occurred in this group because of its size, while relative risks increased stepwise with higher scores. Deviations in heart rate, respiratory rate, and oxygen saturation were most frequent among patients requiring ICU admission or experiencing early mortality, whereas early hospital readmission was most common in patients with low NEWS values (0-4). Together, these findings underscore a core challenge for ED decision support, namely the need to improve discrimination within the large low to intermediate risk population without creating excessive false positives, while also reliably identifying the smaller fraction at highest imminent risk.

### Depth over volume: an ED-first longitudinal acute care dataset

Compared with widely used acute, emergency, and critical care datasets, this resource is distinguished by its ED-first design and breadth of data modalities. Whereas resources such as MIMIC-IV-ED and the Dutch emergency department quality registry NEED include larger numbers of cases with primarily structured clinical variables and short-term in-hospital outcomes [6,24], Acutelines integrates pre-hospital information, detailed ED process and monitoring data, electrophysiological waveforms, laboratory measurements, diagnoses, and long-term outcomes after discharge that include patient-reported outcome measures and mortality. A close comparison can be made with the MC-MED dataset, which also includes waveforms but lacks long-term follow-up [7]. It could serve as a relevant complementary dataset, for example, for external validation. The longitudinal linkage in Acutelines captures the full trajectory from pre-existing health through acute illness to long-term outcomes, enabling identification of risk and protective factors that extend beyond the index admission. Acute presentations often represent decompensation on top of chronic disease, with consequences that extend well beyond the index admission and substantially affect long-term health, functional status, and survival. The addition of prospectively collected biomaterials at ED triage and later during hospitalization if indicated, further supports translational research. Although smaller in size than some national and international databases, Acutelines offers high per-patient data depth and temporal resolution, supporting multimodal, longitudinal, and clinically relevant research across the emergency care pathway.

### Real-world ED data: challenging, but a translational advantage

Equally important is the real-world nature of the data captured within Acutelines. The cohort prioritizes routinely collected, clinically interpretable variables, including demographics, early warning scores, vital signs, laboratory results, and diagnostic codes, embedded within everyday emergency department workflows rather than research-only measurements. Although real-world data are characterized by measurement conditional on clinical indication and non-random missingness, this reflects actual practice and is a key advantage for translation. Where scientific refinement is required, routine data are complemented by de novo data and biomaterial collection, manual curation, and structured adjudication by trained research staff. By closely mirroring routine care, attrition between model development and clinical implementation is reduced, and resulting models are more likely to generalize and integrate into clinical decision making [25].

### Moving beyond one-size-fits-all risk models

The combination of structured triage data, heterogeneous laboratory measurements, time-resolved clinical events, and electrophysiological waveforms provides a strong foundation for advanced data science. Marked heterogeneity in demographics, triage acuity, diagnoses, and outcomes across demographic and clinical subgroups highlights the limitations of one-size-fits-all risk models. Acutelines enables systematic exploration of effect modification and disparities, supporting stratified analyses, prognostic and predictive enrichment, and hypothesis generation across patient subpopulations [26]. Together, the combination of high-resolution real-world data from a largely unselected cohort supports early risk prediction, outcome forecasting across multiple time horizons, missingness-aware modeling, and subgroup-specific performance evaluation, enabling algorithms that move acute care beyond one-size-fits-all decision making.

### Limitations

Several limitations should be considered. First, Acutelines is currently a single-center cohort, which may limit generalizability. This is partly mitigated by the UMCG serving a large and heterogeneous catchment area, including both urban and rural populations and referrals spanning general hospital and tertiary care. In the Netherlands, ED access occurs through referral by a general practitioner or via ambulance, rather than walk-in presentation. This structural difference may influence international comparisons. Moreover, the infrastructure is explicitly designed for external validation and multicenter collaboration, with harmonized data elements, biomaterial collection, and sampling timelines aligned with international partners. Second, laboratory and physiological measurements are obtained selectively based on clinical indication, resulting in variable availability and non-random missingness. While this complicates analysis, it reflects real-world emergency department practice and is advantageous for developing and evaluating models intended for routine clinical deployment; the availability of time-resolved data, rich metadata, and standardized post hoc processing enables transparent handling of missingness and sensitivity analyses. Third, diagnoses coded by clinical staff using SNOMED-CT and ICD-10 are administrative and may not fully capture diagnostic uncertainty or complexity at presentation. This is mitigated by the availability of complementary clinical context, including presenting symptoms, vital signs, laboratory results, electrophysiological waveforms, and longitudinal outcomes, as well as additional scientific adjudication by trained research staff, allowing analyses that extend beyond single administrative labels and better reflect true clinical trajectories.

### Future perspectives

Acutelines is designed as a collaborative infrastructure for the development, testing, validation, and implementation of data-driven models and diagnostic tests in acute care and to serve as a platform to support prospective and interventional studies. Recent work has shown that foundation models trained on even a single physiological modality can generalize across a wide range of clinical prediction tasks beyond traditional diagnostics [12,15,27]. The multimodal and longitudinal structure of Acutelines, integrating triage data, physiological measurements, laboratory results, electrophysiological waveforms, outcomes, and biomaterials, provides a complementary environment to extend, validate, and benchmark such approaches across diverse acute disease states and physiological systems [28,29]. A key objective is to establish Acutelines as an (inter)nationally recognized infrastructure for prospective interventional research, including evaluation of smart algorithms, biomarker-based diagnostics, and clinical trials embedded in routine acute care. Ongoing efforts aim to enable real-time availability of all data streams, including high-resolution electrophysiological waveforms, to support prospective validation and clinical implementation of decision support tools. Furthermore, additional collection of pre-hospital helicopter emergency medical services, ambulance, and general practitioner datastreams and waveforms is a key element in expanding the real-world clinical dataset and enhancing longitudinal data capture. Through cross-institutional and industry collaborations, Acutelines emphasizes knowledge utilization by enabling co-development, validation, certification, and dissemination of tools derived from the infrastructure. Together, these efforts support reproducible research, benchmarking, and validation of clinical AI, and healthcare innovation aligned with learning health system principles.

## Declarations

### Ethics approval and consent

Acutelines and this specific study are approved by the institutional review board (CTC) of the UMCG (#201900635 and #23514). Acutelines is registered at ClinicalTrials.gov under trial registration number NCT04615065. Participants were asked for written informed consent, when applicable by proxy or opt-out.

### Data availability

Although data and biomaterials are not publicly accessible due to privacy and ethical constraints, Acutelines actively supports collaborative research through regulated access procedures, which can be initiated by an email to (for internal and external collaborators) or submitting a study protocol via www.acutelines.nl (for UMCG employees).

### Author contributions (CRediT statement)

Conceptualization: RJW, JMLA, ADS, JL, JCM, EA, NS, HRB. Data curation: RJW, ADS, STH, MAV, AJB, JP. Formal analysis: JMLA, RJW. Funding acquisition: JCM, EA, HRB. Investigation: RJW, JMLA, ADS, JL, STH, MAV, AME, AJB, JP. Methodology: RJW, JMLA, ADS, JL, NS, HRB. Project administration: MAV, AME, JCM, EA, NS, HRB. Resources: JCM, EA, NS, HRB. Supervision: JCM, EA, NS, HRB. Validation: NS, HRB. Visualization: RJW, JMLA. Writing – original draft: RJW, JMLA. Writing – review & editing: RJW, JMLA, ADS, JL, STH, MAV, AME, AJB, JP, JCM, EA, NS, HRB.

## Acknowledgements

The authors thank all participants for contributing their data and biomaterials to Acutelines, as well as the research assistants, team captains, and data managers for their dedicated efforts in data and biomaterial collection. We also acknowledge the UMCG research IT department for facilitating and maintaining the research data infrastructure, as well as the UMCG, in particular the departments of Internal Medicine and Acute Care, for financially supporting the infrastructure.

