## Supplementary material for "Acutelines: a Dutch emergency department-based data-biobank for multimodal, longitudinal acute care research"

### A Individual distributions

#### A.1 Biomaterials

Table A.1: Distribution of biomaterial sample counts per patient visit. Values are reported as timepoint after ED admission, sample and additive type, and n (%) being the number of patient visits with at least one sample of the specific type taken. Note that biomaterials are only taken for a subset of patients in the Reuse+DeNovo inclusion arm (see Methods section), and percentages are therefore relative to the total number of patients eligible for biomaterials.

| Timepoint | Sample type | Additive | Visits n (%) |
|---|---|---|---|
| **0–3 hours** | Serum | Silica | 2207 (63.9) |
| | Plasma | Citrate | 2244 (65.0) |
| | Plasma | EDTA | 2250 (65.2) |
| | Buffycoat | EDTA | 2217 (64.2) |
| | Plasma | Li-Hep | 2029 (58.8) |
| | Wholeblood | PAXgene | 1772 (51.3) |
| | Urine | n/a | 146 (4.2) |
| | Feces | n/a | 23 (0.7) |
| **3–8 hours** | Serum | Silica | 335 (11.6) |
| | Plasma | Citrate | 348 (12.0) |
| | Plasma | EDTA | 352 (12.2) |
| | Buffycoat | EDTA | 343 (11.9) |
| | Plasma | Li-Hep | 310 (10.7) |
| | Wholeblood | PAXgene | 269 (9.3) |
| | Urine | n/a | 152 (5.3) |
| | Feces | n/a | 19 (0.7) |
| **1–2 days** | Serum | Silica | 1161 (40.1) |
| | Plasma | Citrate | 1166 (40.3) |
| | Plasma | EDTA | 1161 (40.1) |
| | Buffycoat | EDTA | 1152 (39.8) |
| | Plasma | Li-Hep | 1053 (36.4) |
| | Wholeblood | PAXgene | 693 (23.9) |
| | Urine | n/a | 537 (18.6) |
| | Feces | n/a | 146 (5.0) |

#### A.2 Waveforms

Table A.2: Summary statistics of available timeseries and waveform data (in minutes) acquired from the Philips bedside patient monitor. Counts show the number of patient visits having at least one measurement and are calculated among non-missing and non-zero observations. Values are reported as median minutes of available data (Q1–Q3).

| **Signal type** | **Sample frequency (Hz)** | **Visits (n)** | **Median Minutes (Q1–Q3)** |
|---|---|---|---|
| Systolic Blood Pressure | variable | 21 916 | 151 (80–296) |
| Diastolic Blood Pressure | variable | 21 915 | 151 (80–296) |
| $SpO_2$ | 1 | 25 657 | 148 (65–274) |
| HR | 1 | 18 629 | 146 (76–234) |
| Respiratory Rate | 1 | 1 089 | 55 (30–90) |
| Respiratory Impedance | 125 | 18 353 | 142 (72–228) |
| PPG | 125 | 26 038 | 157 (76–281) |
| ECG rhythm | variable | 23 586 | 165 (90–282) |
| ECG lead I | 500 | 18 624 | 142 (72–229) |
| ECG lead II | 500 | 18 684 | 144 (74–232) |
| ECG lead V1 | 500 | 8 734 | 131 (60–217) |
| ECG lead V5 | 500 | 4 718 | 34.5 (14–134) |
| Capnography | 125 | 870 | 49 (25–74) |

#### A.3 Demographics

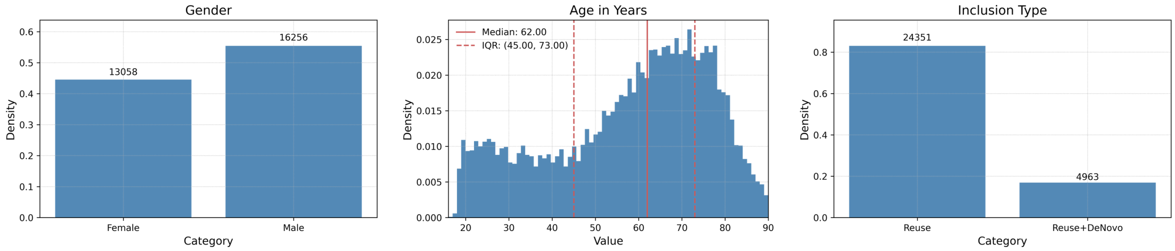


Figure A.3: Demographics

#### A.4 Comorbidities

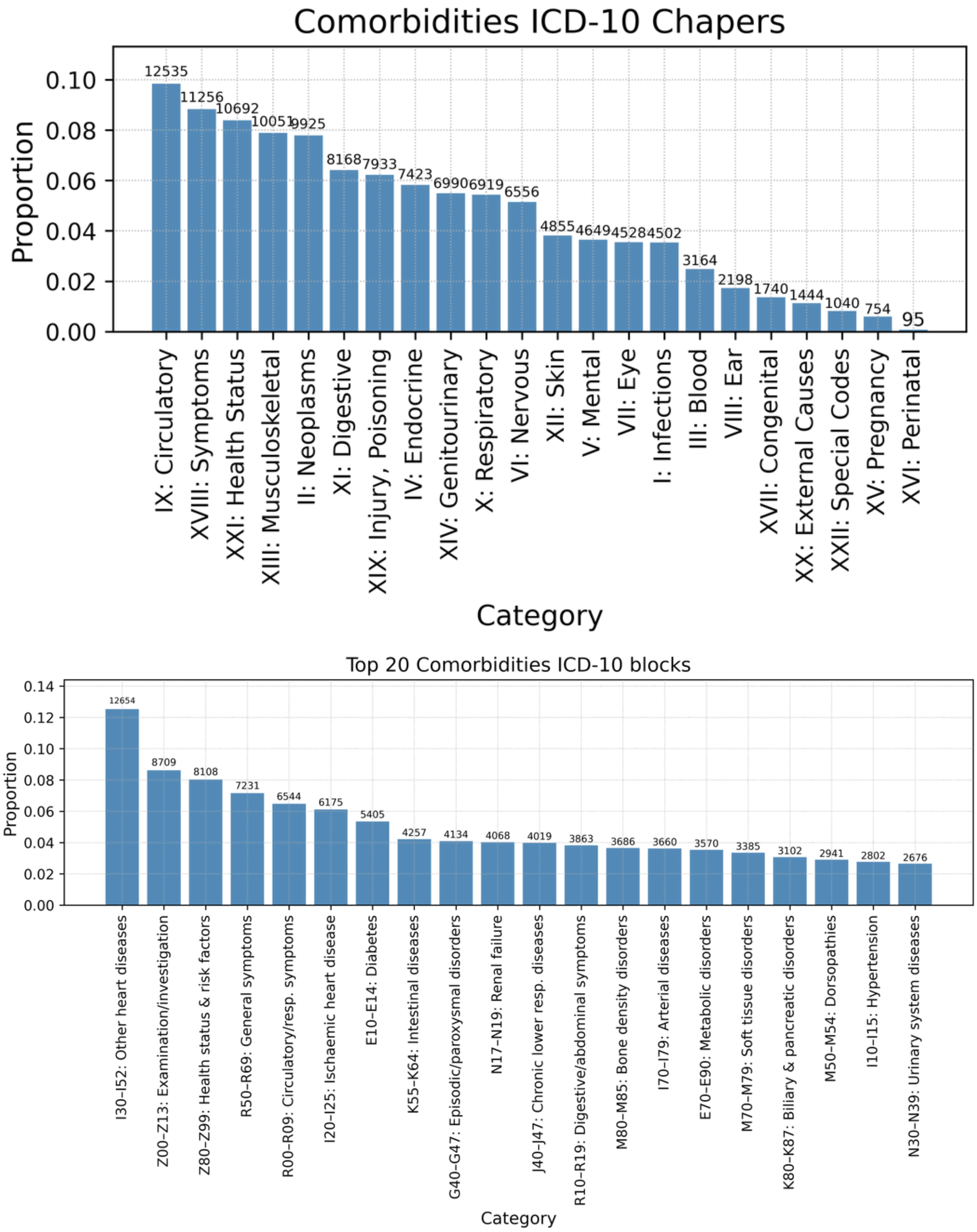


Figure A.4: Comorbidities

#### A.5 Diagnoses

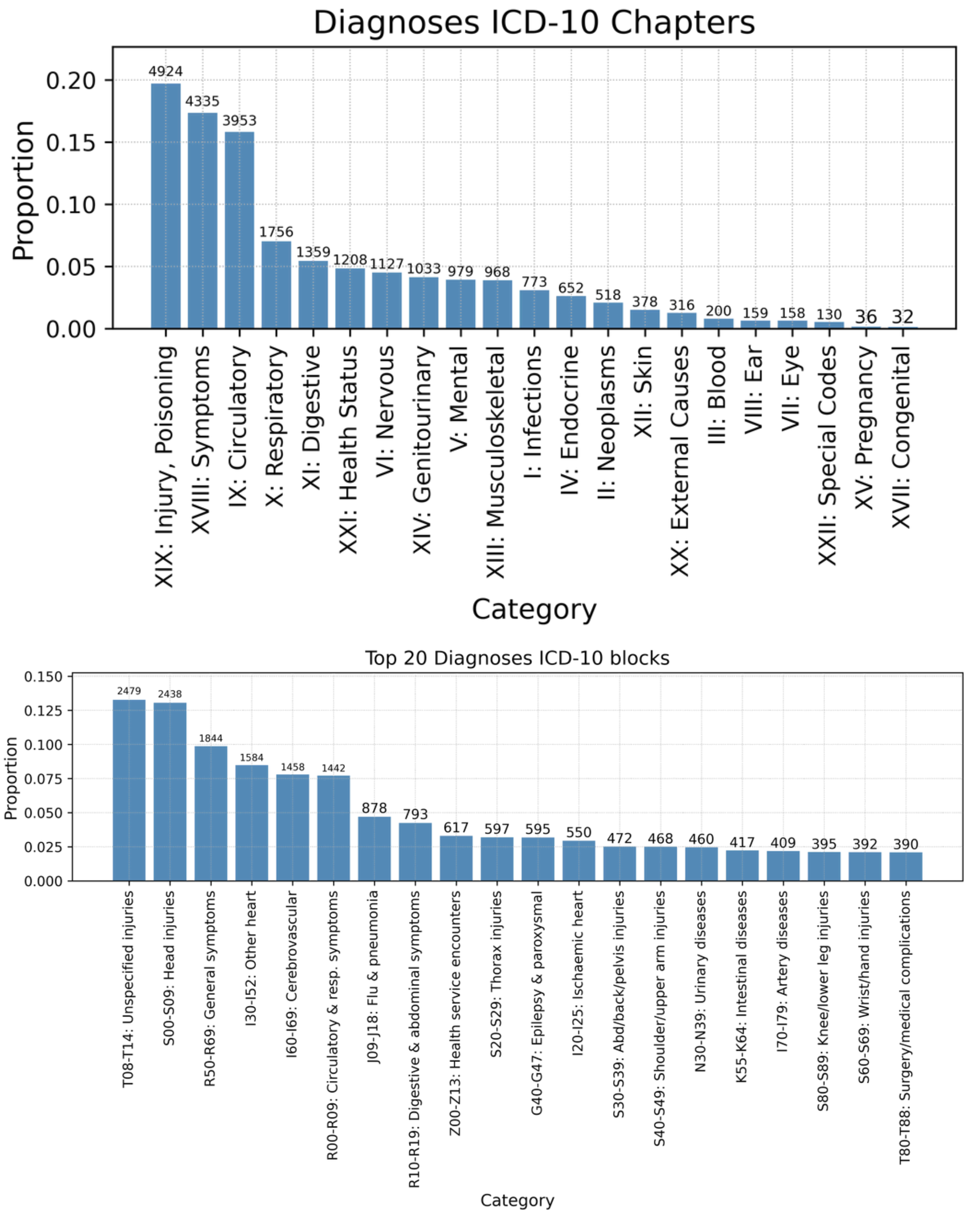


Figure A.5: Diagnoses

#### A.6 Vital parameters

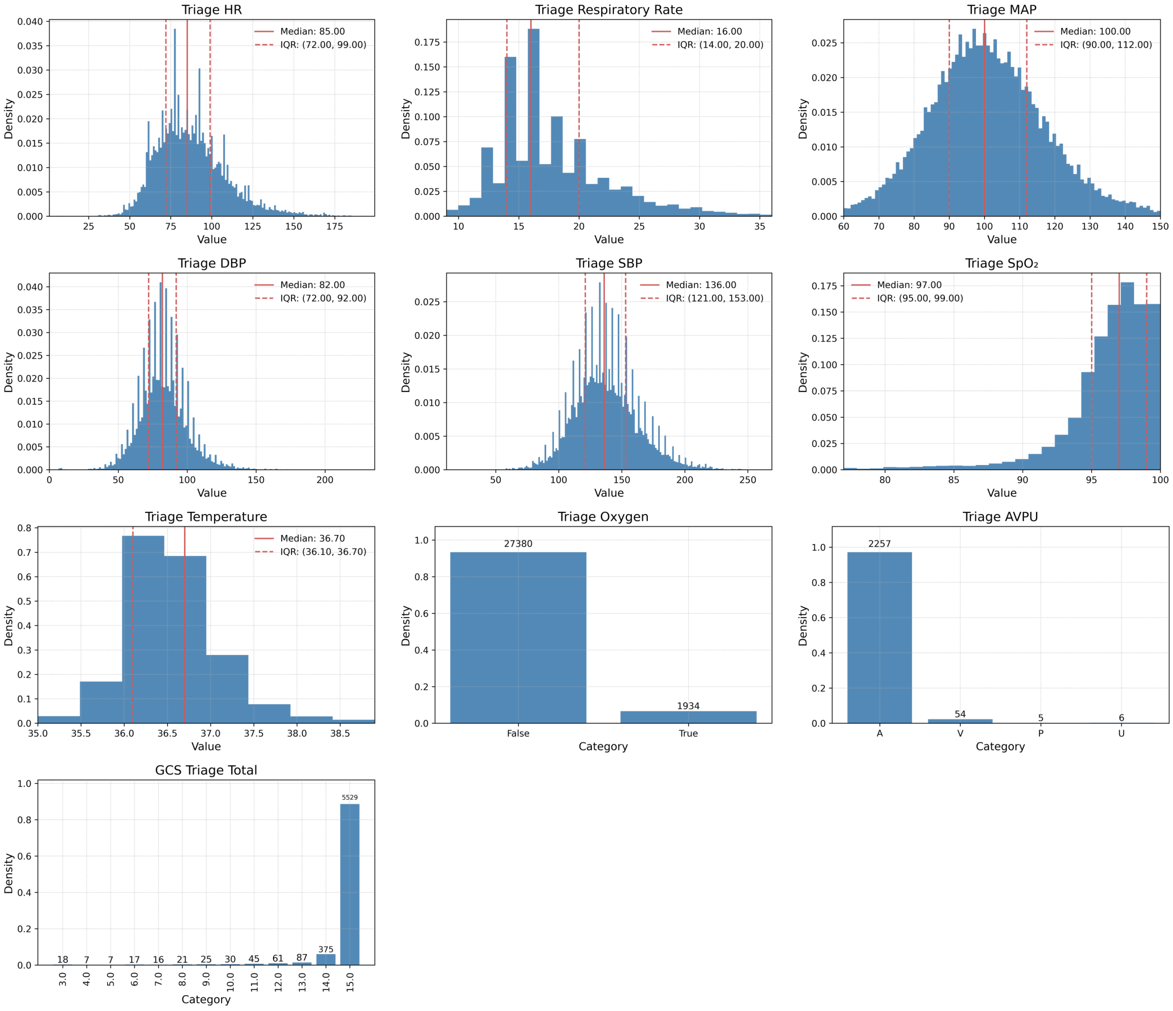


Figure A.6: Vital parameters

#### A.7 Early Warning Scores

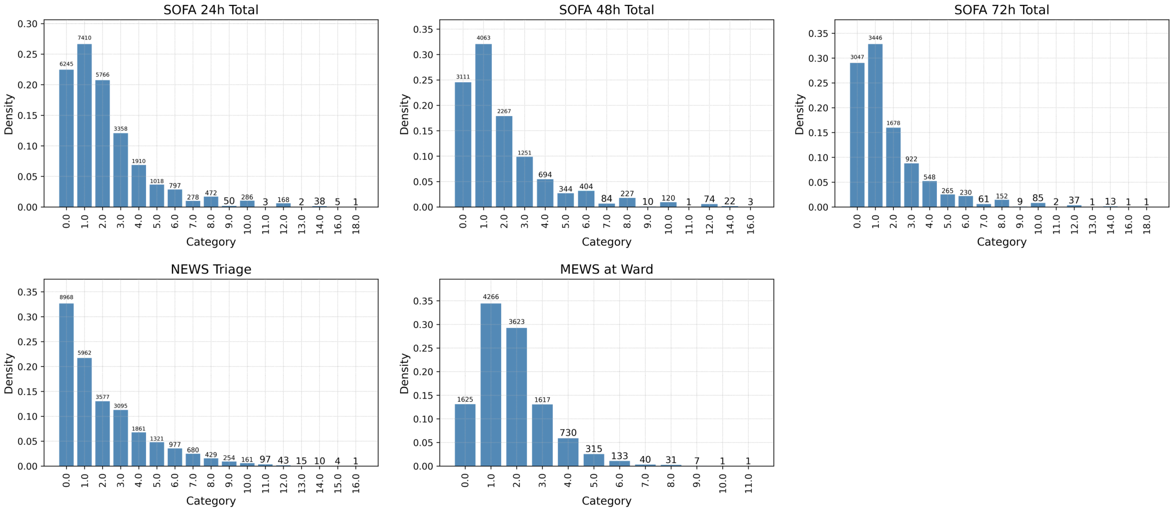


Figure A.7: Early Warning Scores

#### A.8 Logistics

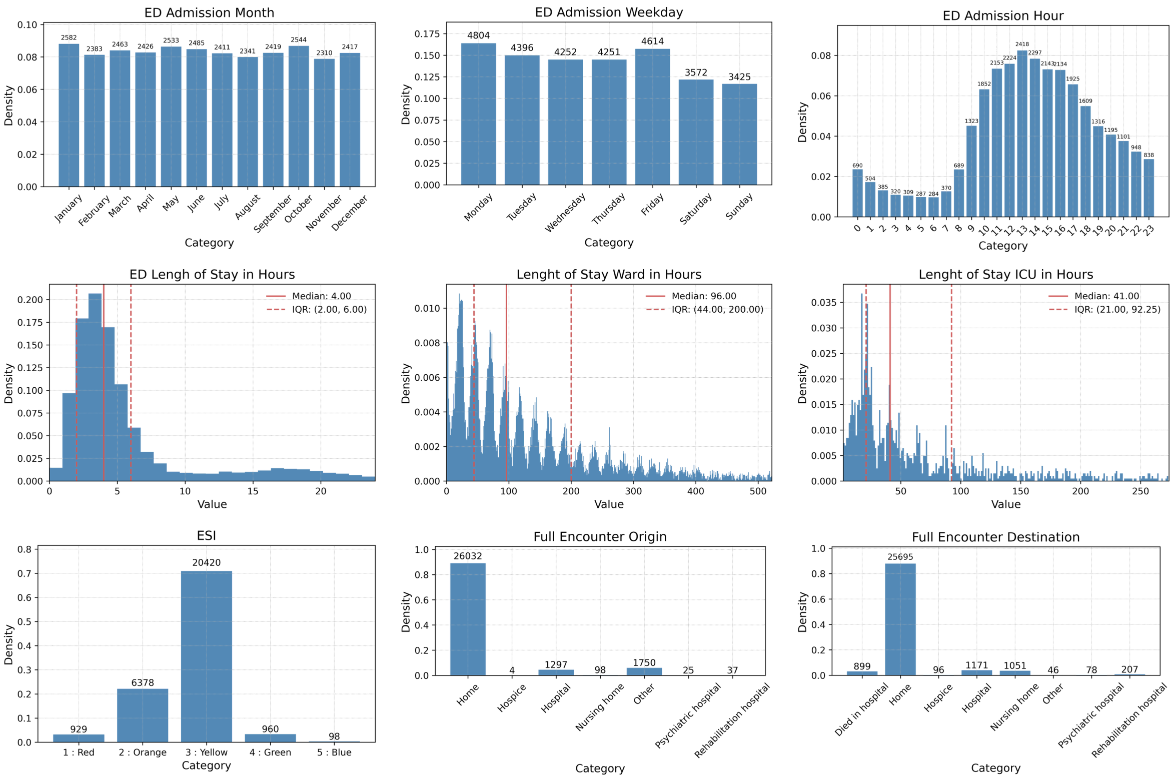


Figure A.8: Logistics

#### A.9 Hematological laboratory values

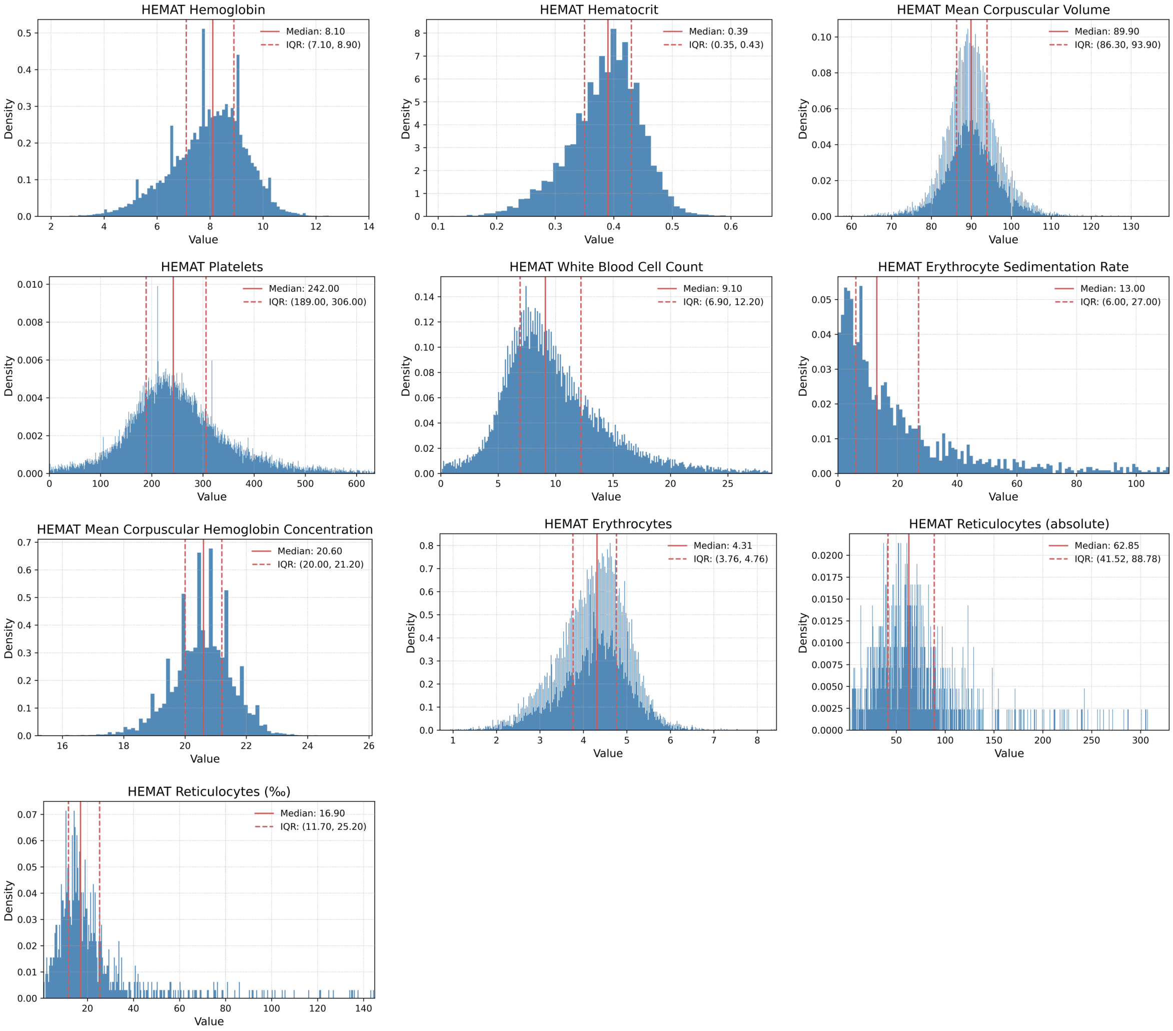


Figure A.9: Hematological laboratory values

#### A.10 White blood cell differentation

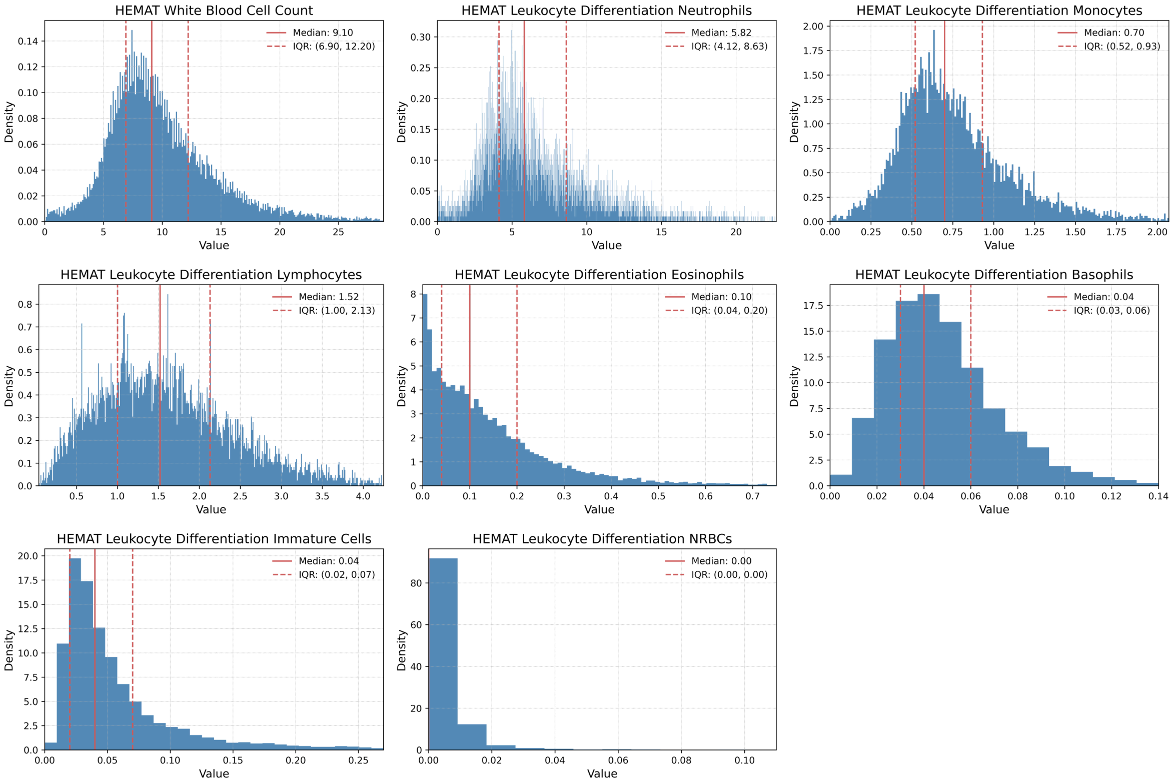


Figure A.10: White blood cell differentation

#### A.11 Chemical laboratory values

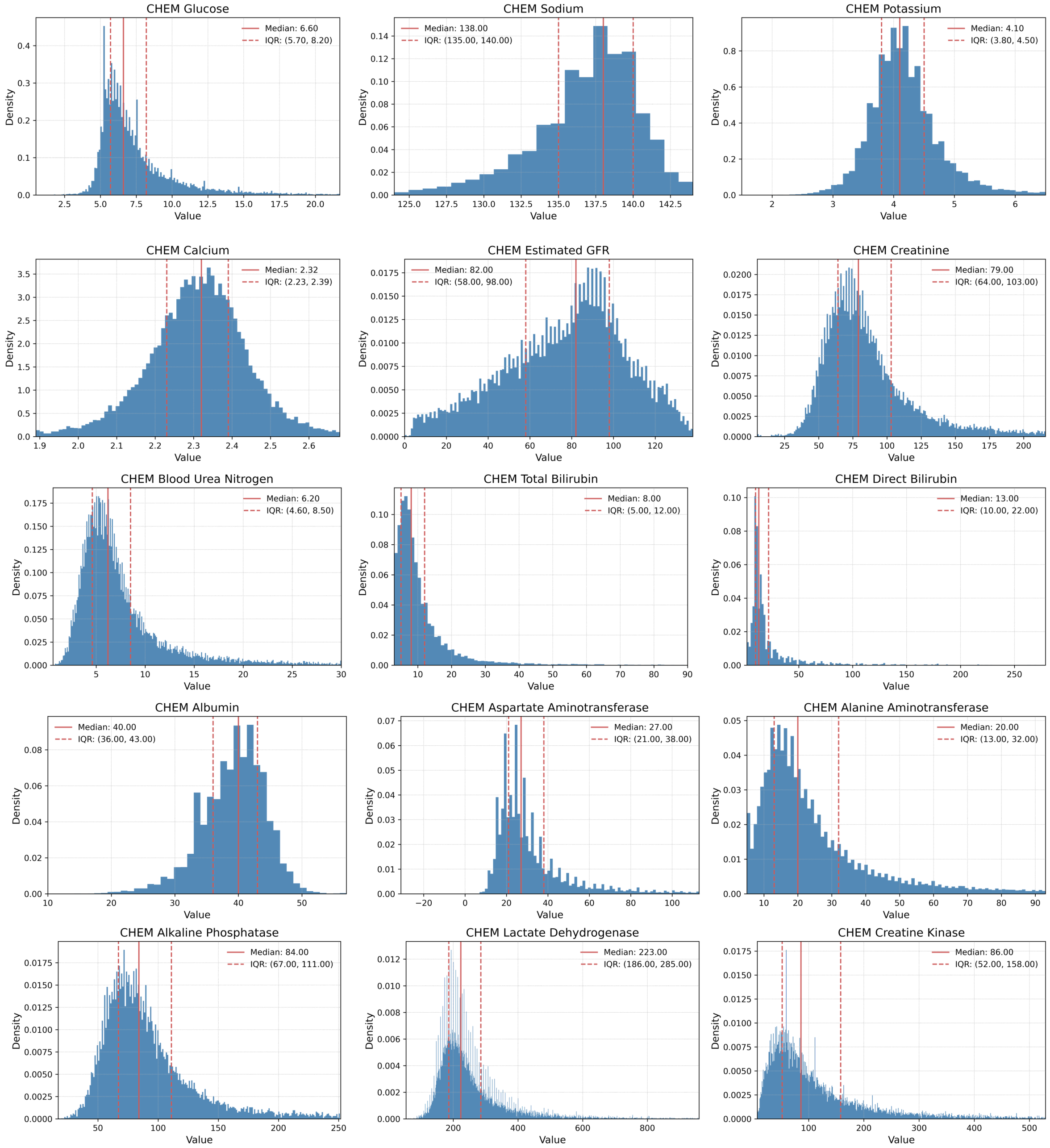


Figure A.11-I: Chemical laboratory values I

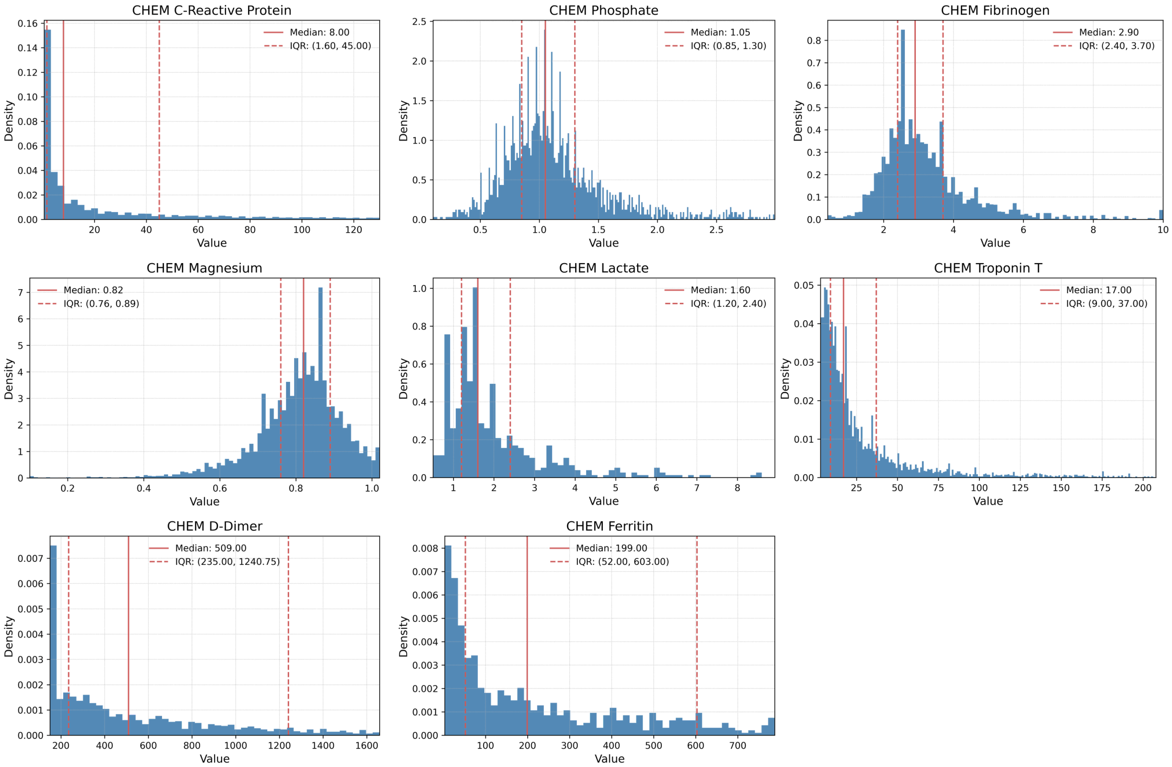


Figure A.11-II: Chemical laboratory values II (continued)

#### A.12 Blood gas analysis

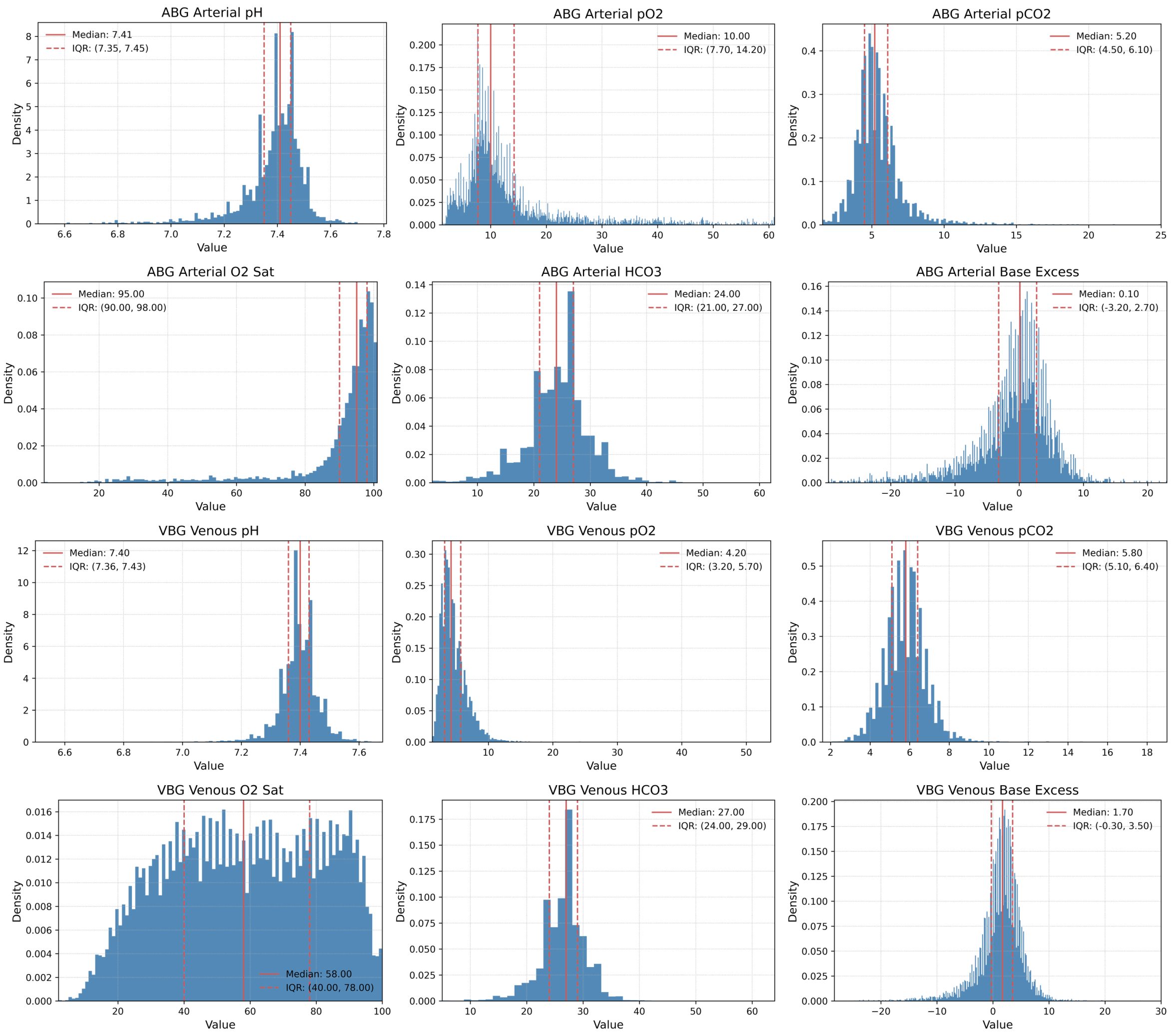


Figure A.12-I: Blood gas analysis I

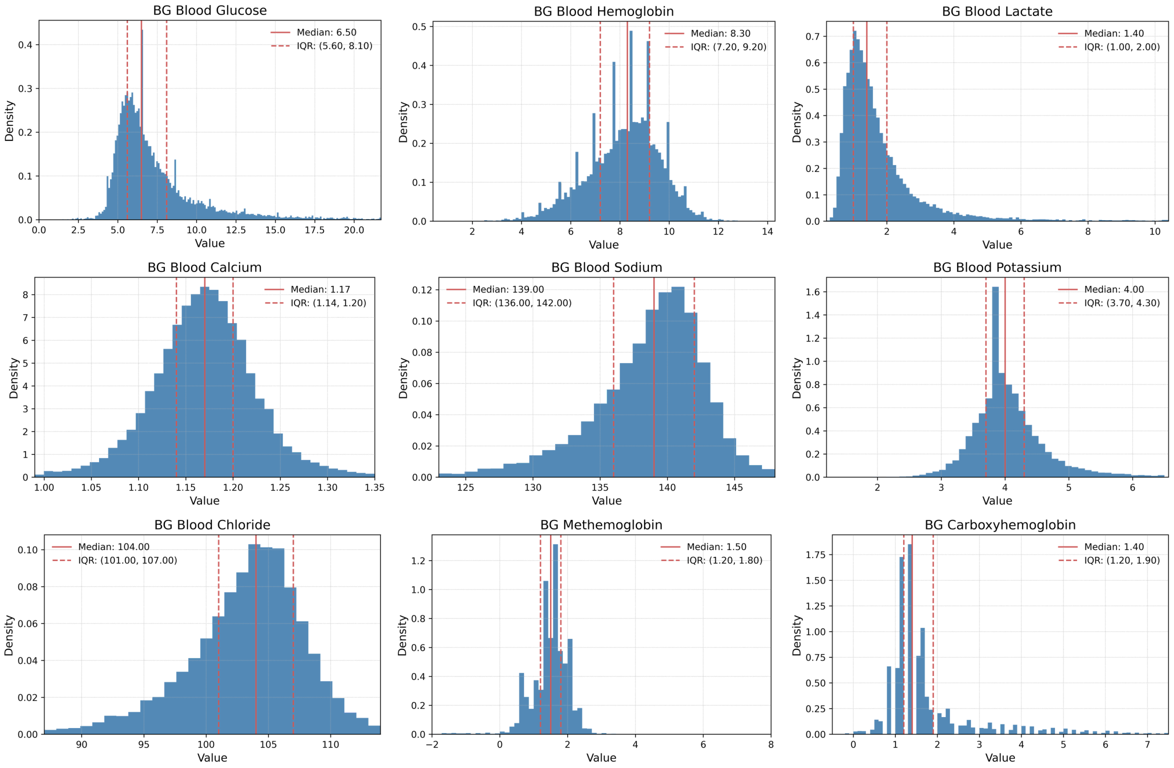


Figure A.12-II: Blood gas analysis II (continued)

#### A.13 Outcome

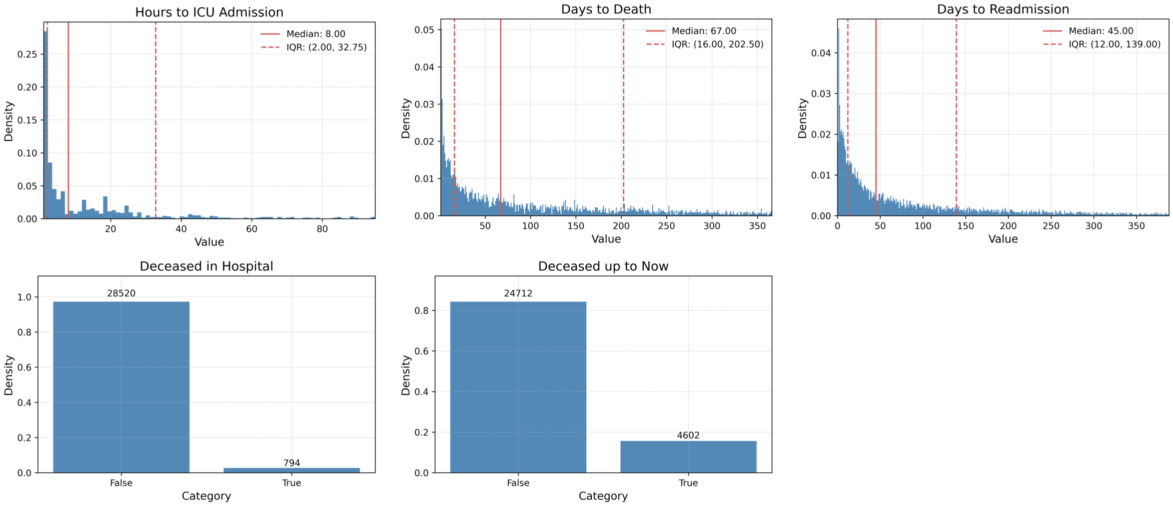


Figure A.13: Outcome

#### A.14 Outcome: time to event Kaplan–Meier curves

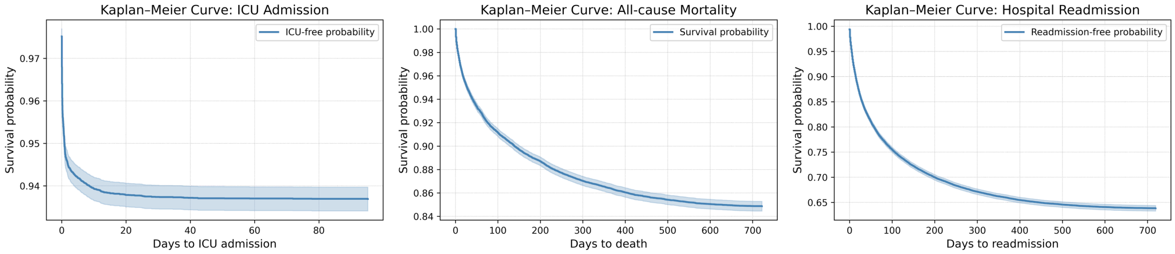


Figure A.14: Outcome Kaplan–Meier curves illustrating the probability of ICU admission, all-cause mortality, or hospital readmission over time.

### B Row-wise heatmaps

#### B.1 ICD-10 clusters

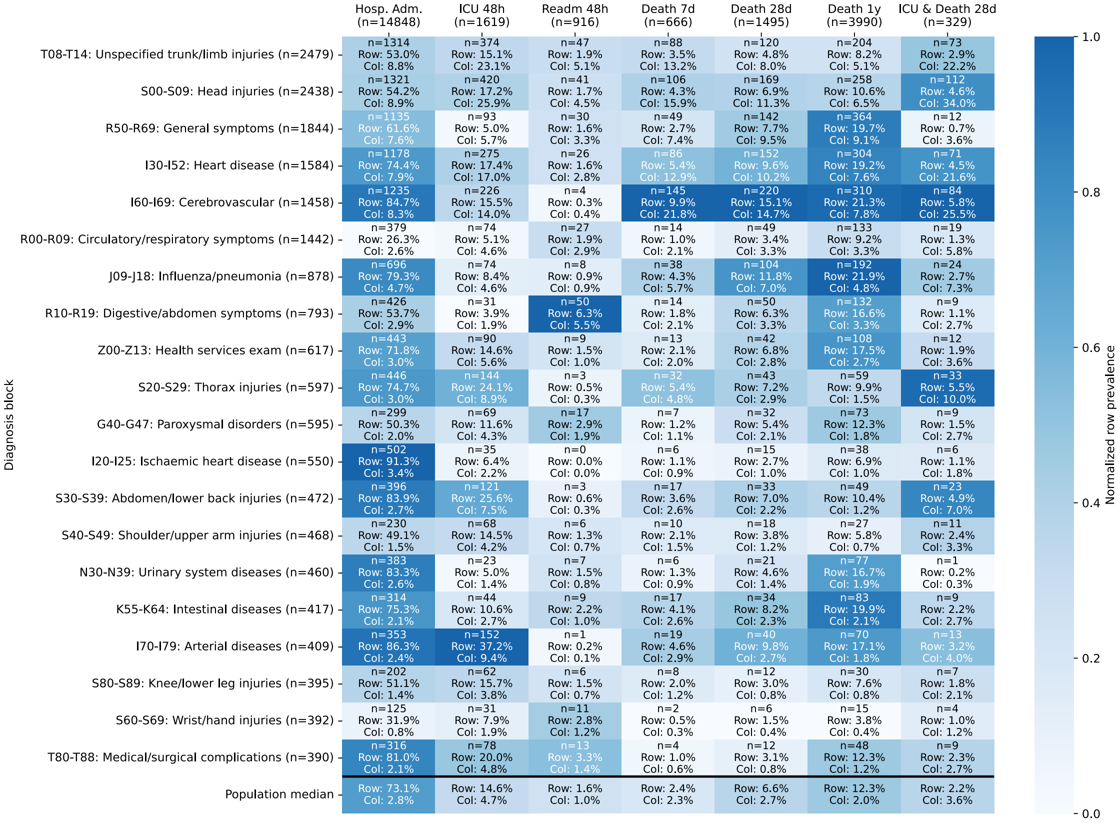


Figure B.1: Top 20 ICD-10 clusters stratified by clinical outcomes relative to emergency department presentation. Outcomes include hospital admission, ICU admission within 48 hours, readmission within 48 hours, and mortality at 7 days, 28 days, 1 year, and ICU or death within 28 days. Each cell reports absolute counts together with row- and column-level percentages. Color intensity reflects row-wise normalized prevalence, with darker shades indicate a higher prevalence of a given outcome relative to other diagnoses within the same outcome. Hosp. Adm.: Hospital Admission; ICU: Intensive Care Unit; Readm: Readmission.

#### B.2 Vitals

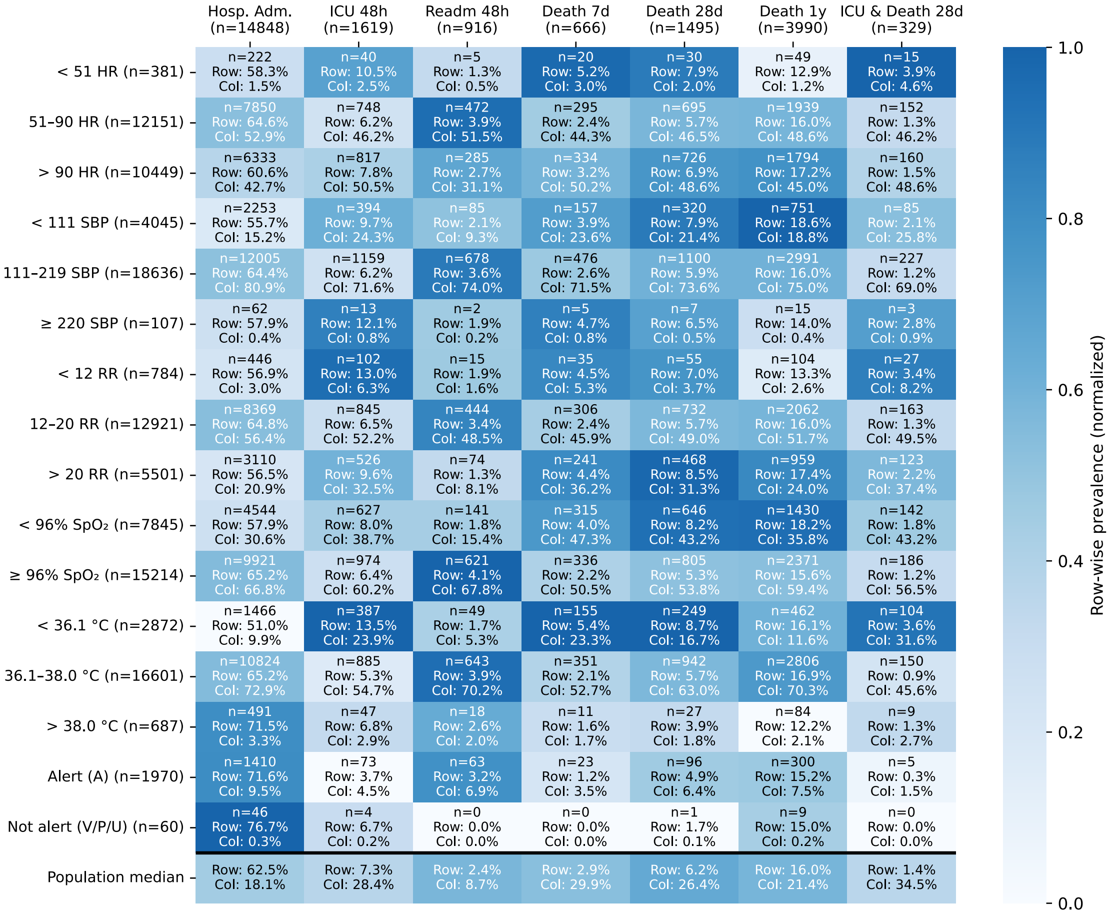


Figure B.2: Vital parameters at triage stratified by clinical outcomes relative to emergency department presentation. Outcomes include hospital admission, ICU admission within 48 hours, readmission within 48 hours, and mortality at 7 days, 28 days, 1 year, and ICU or death within 28 days. Each cell displays both row- and column-level prevalence. Color intensity reflects row-wise normalized prevalence, with darker shades indicating a higher prevalence of a given outcome, relative to other vital parameter categories within the same outcome. Hosp. Adm.: Hospital Admission; ICU: Intensive Care Unit; Readm: Readmission; HR: Heart rate; RR: respiratory rate; $SpO_2$: peripheral oxygen saturation; SBP: Systolic Blood Pressure; °C: degree Celsius; A: Alert; V: Verbal; P: Pain; U: Unresponsive.

#### B.3 NEWS

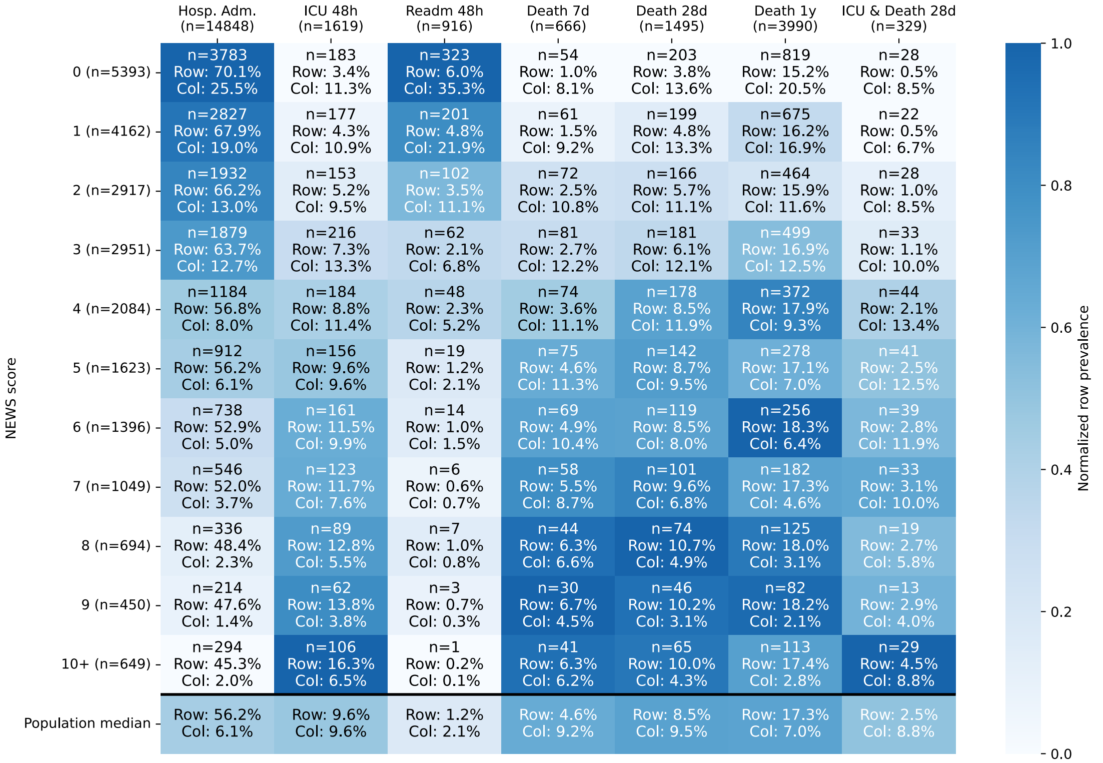


Figure B.3: National Early Warning Score (NEWS) at triage stratified by clinical outcomes relative to emergency department presentation. Outcomes include hospital admission, ICU admission within 48 hours, readmission within 48 hours, and mortality at 7 days, 28 days, 1 year, and ICU or death within 28 days. Each cell displays both row- and column-level prevalence. Color intensity reflects row-wise normalized prevalence, with darker shades indicating a higher prevalence of a given outcome relative to other NEWS scores/categories within the same outcome. Hosp. Adm.: Hospital Admission; ICU: Intensive Care Unit; Readm: Readmission; NEWS: National Early Warning Score.
